# The impact of non-pharmaceutical interventions on the prevention and control of COVID-19 in New York City

**DOI:** 10.1101/2020.12.01.20242347

**Authors:** Jiannan Yang, Qingpeng Zhang, Zhidong Cao, Jianxi Gao, Dirk Pfeiffer, Lu Zhong, Daniel Dajun Zeng

## Abstract

The emergence of coronavirus disease 2019 (COVID-19) has infected more than 37 million people worldwide. The control responses varied across countries with different outcomes in terms of epidemic size and social disruption. In this study, we presented an age-specific susceptible-exposed-infected-recovery-death model that considers the unique characteristics of COVID-19 to examine the effectiveness of various non-pharmaceutical interventions (NPIs) in New York City (NYC). Numerical experiments from our model show that the control policies implemented in NYC reduced the number of infections by 72% (IQR 53-95), and the number of deceased cases by 76% (IQR 58-96) by the end of 2020, respectively. Among all the NPIs, social distancing for the entire population and the protection for the elderly in the public facilities is the most effective control measure in reducing severe infections and deceased cases. School closure policy may not work as effectively as one might expect in terms of reducing the number of deceased cases. Our simulation results provide novel insights into the city-specific implementation of NPIs with minimal social disruption considering the locations and population characteristics.

## Introduction

The outbreak of coronavirus disease 2019 (COVID-19) has become a global pandemic with unanticipated consequences to the global community. As of 13 October 2020, severe acute syndrome coronavirus 2 (SARS-CoV-2), the cause of COVID-19, has infected more than 37 million people and resulted in more than 1 million deaths (1). A variety of non-pharmaceutical interventions (NPIs) were introduced to reduce the transmission by lowering contact intensity at different locations (2), such as school closure, workplace shutdown, and the closure of bars, churches, and other public facilities, which has been shown to be successful in China (3), South Korea (4), and other countries (5). The NPIs, such as city-wide school closures, were implemented as part of its State of Emergency plan (6) in New York City (NYC) that has been identified as a major epicenter with over 258,000 cases and 23,915 confirmed deaths (7). However, the effectiveness of these NPIs remains unclear, promoting a critical need to evaluate them and to derive more effective NPIs with the consideration of social disruption.

Increasing evidence shows that the demographic structure and age-specific contacts at different locations play an essential role in the COVID-19 epidemic, as well as the effectiveness of varying NPIs (8-12). For instance, school closure is useful in controlling the infections among the young people (13, 14), who tend to have mild or moderate symptoms and usually can recover without treatment. On the other hand, severe and deceased cases are often among the elderly and those with comorbidities (15). According to the data from the CDC of the US, 79.2% of death in the US are the population whose age is over 64. (16) Existing studies (17, 18) addressed this problem by dividing the population into several age groups and defining the social contacts at different locations. However, utilizing these unique epidemiological characteristics to reduce the total number of infections and the severe and deceased cases and the social disruption is under-researched.

In this article, we develop a novel age-specific susceptible-exposed-infected-recovery-death (A-SEIRD) model (19, 20) to examine the effect of a set of common NPIs on reducing the total number of infections and the severe and deceased cases. More importantly, we obtain the NPIs that can contain the epidemic with minimal social disruption by quantitatively examining all possible NPIs. The latest epidemiological parameters (21, 22) of COVID-19 were adopted by the A-SEIRD model.

## Methods

### Adjusted Contact Matrix and Transmission Rate

The contact patterns vary across locations. Previous studies (17, 18) divide the contact intensity into four locations within a city: households, workplaces, schools, and public facilities represented as *C*^*H*^, *C*^*W*^, *C*^*S*^, and *C*^*P*^ individually. Note here the original contact matrix *C* is estimated by survey data achieved in 2008 (17). To get the updated contact matrix, we adjust the original contact matrix *C* by the latest population structure that refers to the age distribution which is assumed to be static during the simulation period for the same city. We divide the total population of a city into 17 age groups including 16 groups with a 5-year band between birth and 80-year-old, and the 17th group representing aged >80. The adjusted contact intensity (*M*_*ij*_) of age group *j* made by age group is obtained by taking the product of *C*_*ij*_ and the ratio of the current population sizes of age group *j* (*p*_*j*_) and age group *j* (*p*_*j*_) as follows:

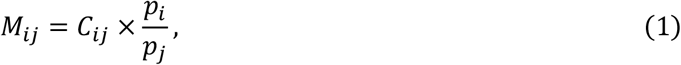

The overall social mixing pattern is defined as the weighted sum of the adjusted contact matrix across the four locations,

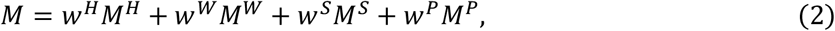

where *w*^*H*^, *w*^*W*^, *w*^*S*^, *w*^*P*^ are the location-specific decay factors compared to the normal situation (*w*^*H*^ = *w*^*W*^ = *w*^*S*^ = *w*^*P*^ = 1) and the effects of different NPIs are reflected by the reduction of these decay factors. The contact matrixes of the four locations under the normal situation are shown in Figure 1.

**Figure 1:**
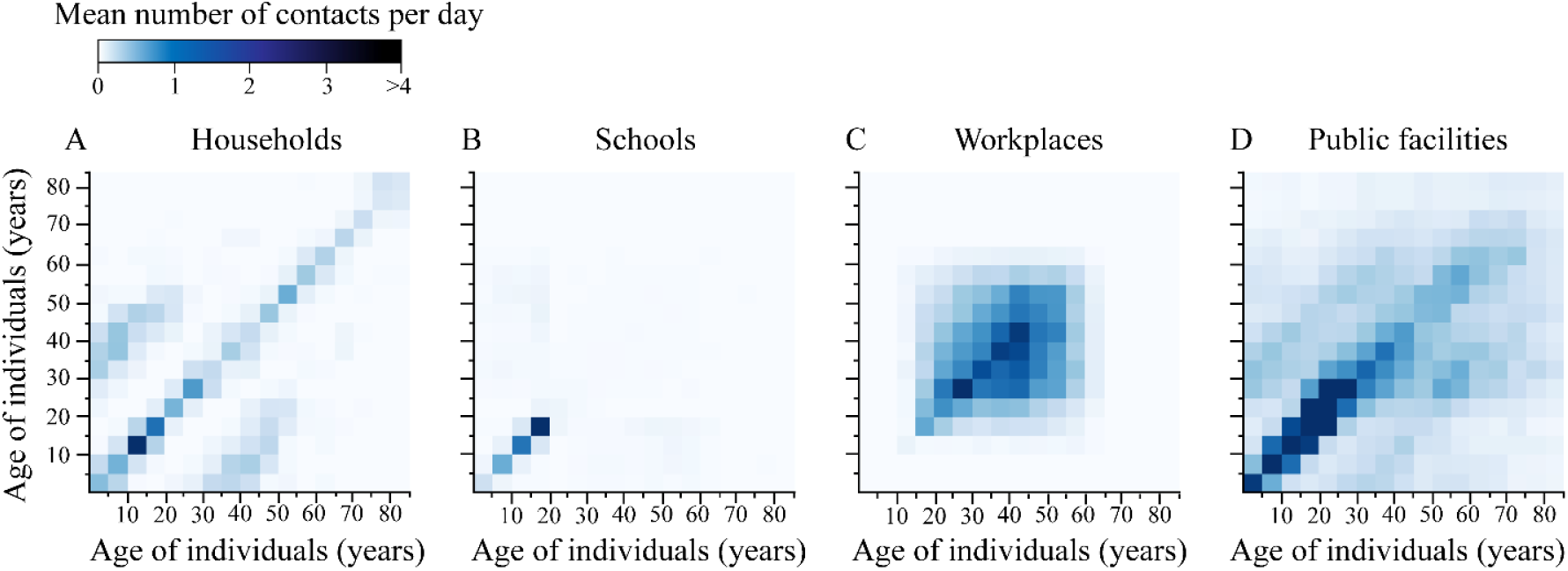
Age-specific and location-specific contact intensities under normal situation for NYC. (A), (B), (C), and (D) denote the contact intensity among all the 17 age groups, respectively. The grid in each panel represents the mean number of contact per day. The colour change denotes the value change: more blue, more contacts.

The transmission rate *β* varies with NPIs shown on the overall adjusted social mixing pattern *M*. Following Prem et al. (17) we projected the adjusted contact matrix with the basic reproduction number to the transmission rate as follows:

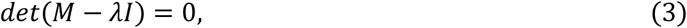

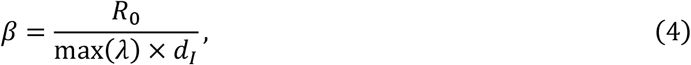

where *R*_0_ is the basic reproduction number and *d*_*I*_ is the average length of the infectious period.

### The A-SEIRD Model

The proposed A-SEIRD model is based on the classic SEIR model (19, 20). In the classic SEIR model, the population is divided into four groups according to the infection status: susceptible (*S*), exposed (*E*), infected (*I*), and recovery (*E*). Based on the evidence of the asymptomatic infections of COVID-19 (23, 24), we add three new statuses for the infected individuals: asymptomatically infected (*I*^*as*^), mildly infected (*I*^*m*^) and severely infected (*I*^*s*^). Susceptible individuals may become exposed after contacting infected ones. Exposed individuals may become either mildly infected or asymptomatically infected. We assume that the deceased cases come from both mild and severe infections, and all asymptomatically infected individuals will recover. The model structure is presented in Figure 2.

**Figure 2:**
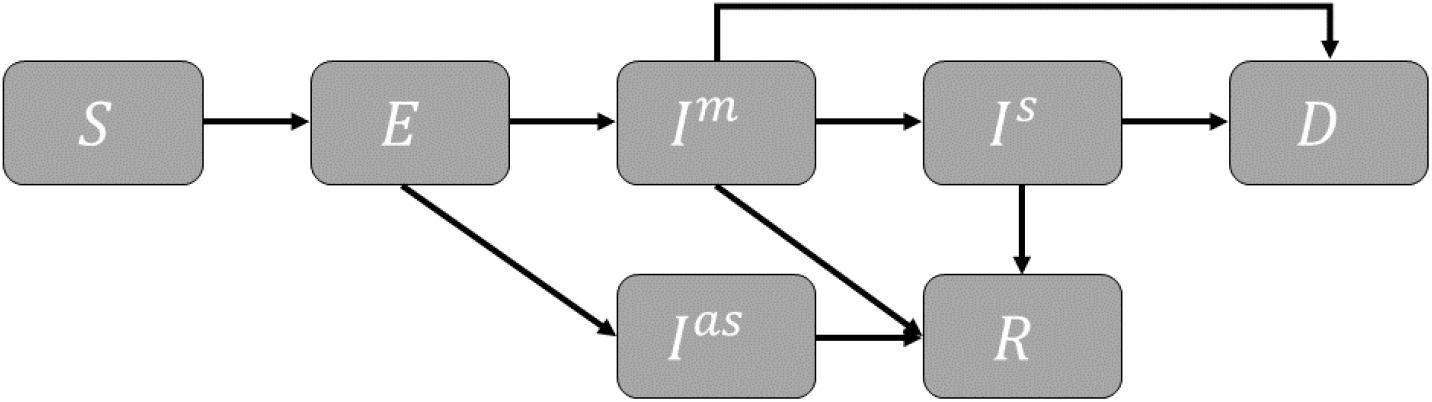
The proposed age-specific SEIRD model.

Considering the huge drop in human movement between cities (25), we assume that NYC is a closed system throughout the course of the epidemic (from 1 Jan 2020 to 31 Dec 2020) and initially there are 1,000 infected individuals for simplicity. The initially infected individuals are distributed in different age groups with the same proportion (0.016%). The epidemic parameters are age-dependent, such as the elder individuals have a higher severe rate and death rate than the youth (22).

For a given age group *i*, the epidemic transitions can be described by

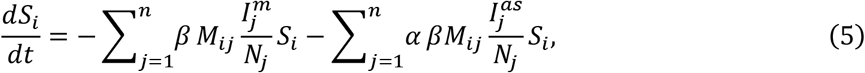

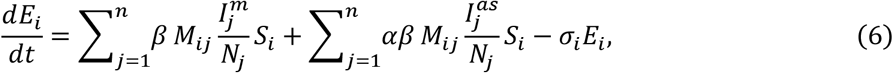

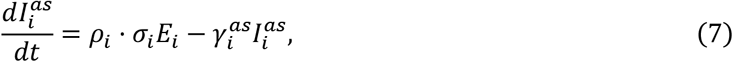

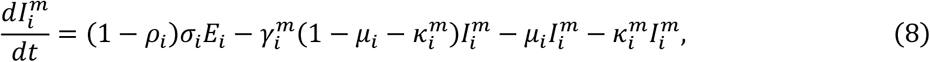

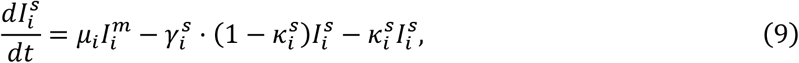

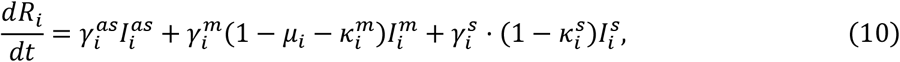

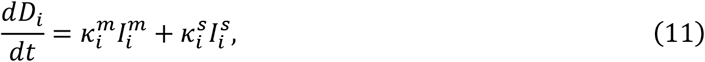

where 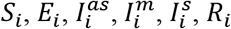, and *D*_*i*_ denote the number of susceptible, latent, asymptomatic infectious, mild infectious, severe infectious, recovery, and deceased individuals at age group *i*, respectively. *N*_*j*_ is the number of individuals in age group *j. β* is the transmission rate that differs by the contact intensity and population structure and *α* is the proportion of the transmission rate of the asymptomatic infections over that of the mild infections (26). Note that here we assume the severe infections will not spread the virus due to their limited mobility (usually isolated in hospital). *M*_*ij*_ denotes the total contact intensity of age group *j* made by age group *i*. For individuals in the age group 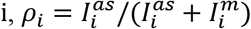 is the estimated fraction of asymptomatic cases among both asymptomatic and mild infections; *σ*_*i*_ = 1/*d*_*L*_ is the daily probability that an exposed individual becomes infectious (either asymptomatic or mild) where *d*_*L*_ is the average latent period; 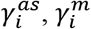, and 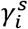 are the daily recovery probability for the asymptomatic, mild, and severe infections, respectively. They are related to specific disease period: 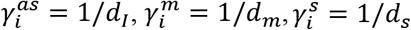 where *d*_*I*_, *d*_*m*_, *d*_*s*_ are the infectious period, mild duration, and severe duration generated from literature, respectively. *μ*_*i*_ is the daily probability of hospitalisation that a mildly infected patient becomes severely infected. 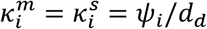 is the daily crude mortality rate, where *ψ*_*i*_ is the crude mortality rate and *d*_*d*_ is the period from the symptom onset to death. The crude mortality rate *ψ*_*i*_ is estimated from the official confirmed case report (27) which equals the proportion of the number of deceased individuals to the infected individuals within one age group. The parameters used in the model from literature are presented in Table 1 and Table 2.

**Table 1.**
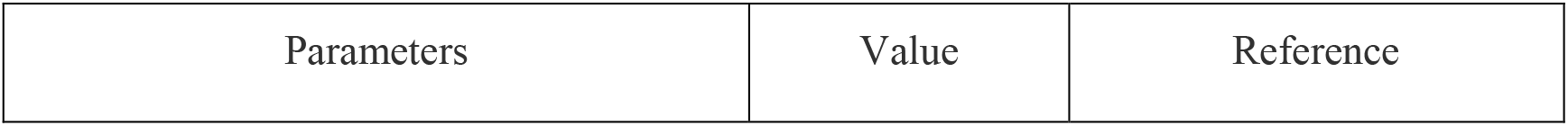
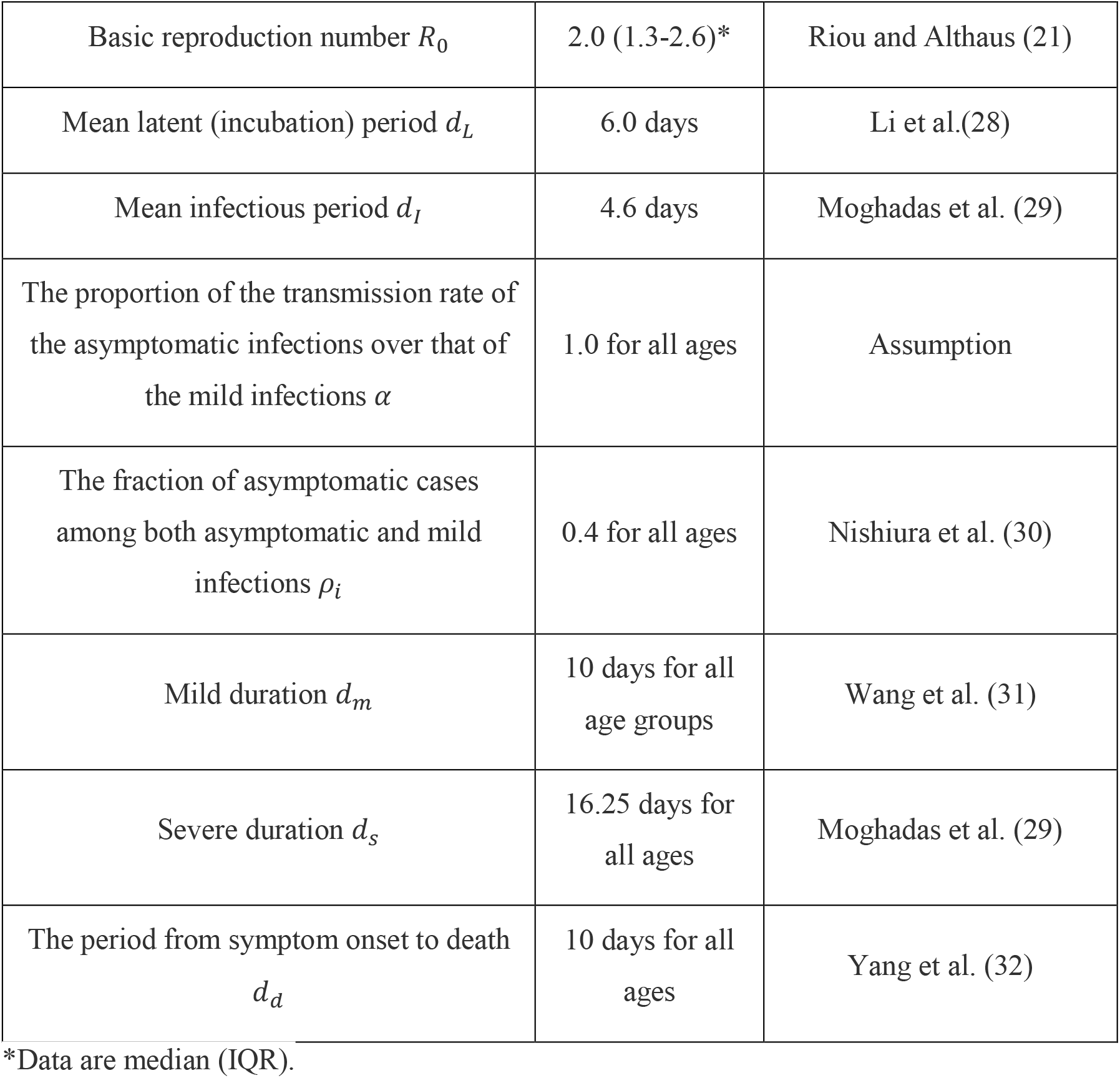
The parameters used in the model from the literature.

**Table 2.** The age-specific parameters used in the model from the literature.

| Description | 0-19 | 19-44 | 45-65 | 65-75 | 75+ | Reference |
| --- | --- | --- | --- | --- | --- | --- |
| Daily probability of hospitalisation<br>$\mu$ | 0.0250 | 0.0320 | 0.0320 | 0.6400 | 0.6400 | Moghadas et al. (29) and WHO-China Joint Mission Report (33) |
| Crude mortality rate $\psi$ | 0.0215 | 0.0782 | 0.2257 | 0.4600 | 0.6085 | Estimated from official confirmed case report (27) |

### Non-Pharmaceutical Intervention Scenarios

Some study (34) points out that the seeding of the virus likely occurred in the US between 25 December 2019 and 15 January 2020, thus we set the starting date of the simulation to be 1 January 2020 for a better fitting, and simulate the epidemic based on the real-time responses by the government. Before the state governor declared a state of emergency in New York State on 7 March, we assume the residents have already had a certain degree of awareness of the COVID-19, and thus the contact intensities at schools, workplaces, and public facilities were reduced by 20%. Between the emergency declaration and the state-wide stay-at-home order (7 March to 22 March), the awareness of precaution has been elevated to reduce the contact intensity at workplaces, schools, and public facilities by 50%. After the state-at-home order was put in place, the city was at a full lockdown status (6), so we set the decay factor of the contact intensity at schools to 0 and further reduce the contact intensities at workplaces and the public facilities to 30% of the normal situation. To model the four reopening phases (35) implemented on 8 June, 22 June, 6 July, and 20 July, respectively, we assume the contact intensities at both the workplaces and public facilities increase by 5% at each phase. In addition, we simulate the scenario in which NYC has a larger-scale reopen back to the situation before the state-at-home order (50% for all locations except household) on 1 September. The decay factor of contacts in households is set to 1 over the course of the epidemic.

We investigate five common NPIs: theoretical no intervention, school closure, social distancing for the entire population, social distancing for the elderly (age > 64 years) (36), and adaptive policy proposed by Ferguson et al. (37). In the adaptive policy, a stringent control measure (full lockdown) is triggered when certain conditions are met. Here we assume that the condition is the number of daily new severely infected cases is over *N*. We examined two values of *N* (100 and 150) for the robustness of results. The details of the decay factor setting for each NPIs are shown in Table 3. We assume the NPIs were implemented through the whole course of the epidemic simulation.

**Table 3.** Summary of NPIs considered.

| NPIs | Description |
| --- | --- |
| Theoretical no intervention | $M = M^H + M^W + M^S + M^P$ |
| School closure | $M = M^H + M^W + 0 \times M^S + M^P$ |
| Social distancing for the entire population | $M = M^H + 0.5M^W + 0.5M^S + 0.5M^P$ |
| The social distancing for the elderly (age > 64 years) | $M = M^H + w^E M^W + w^E M^S + w^E M^P$ $w^E = \begin{bmatrix} 1 & 0 & 0 & \cdots & 0 & 0 & 0 \\ 0 & 1 & 0 & \cdots & 0 & 0 & 0 \\ 0 & 0 & \vdots & \cdots & \vdots & 0 & 0 \\ 0 & 0 & 0 & \cdots & 0 & \vdots & 0 \\ 0 & 0 & 0 & \cdots & 0.5 & 0 & \vdots \\ \vdots & \vdots & \vdots & \cdots & 0 & 0.5 & 0 \\ 0 & 0 & 0 & \cdots & 0 & 0 & 0.5 \end{bmatrix}$ |
| Adaptive policy | $M = M^H + 0 \times M^W + 0 \times M^S + 0.1M^P$<br>*if* $N_s > 100$ (150) where $N_s$ denotes the number of new severe infections per day |

### Identifying Effective NPIs Considering Social Disruption

The objective of NPIs should not be limited to reducing infections without the consideration of social disruption. First, we measure the social disruption caused by the NPIs at the four locations. Here, the social disruption at a location is measured as (1 − *w*)*M* × *s*, which is the product of the reduced amount of contacts (1 − *w*)*M* and the relative importance of these contacts *s*. Thus, the total social disruption *L* caused by the NPIs can be represented as follows,

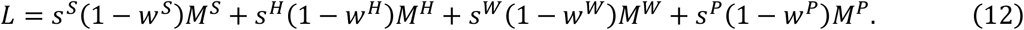

Second, we aim to identify the NPIs that reduce the social disruption *L* as much as possible under the condition that the COVID-19 epidemic does not exceed that of the actual NPIs implemented in NYC. Specifically, we consider three measures of the COVID-19 epidemic: the number of all infections, the number of severe infections, and the number of deceased cases as of 31 December 2020, respectively. It is challenging to quantify the relative importance of the activities at different locations in a very detailed manner. In this study, we make the following intuitive assumptions about relative importance: workplaces have the highest importance, *s*^*W*^ = 1; public facilities has the second highest importance, *s*^*P*^ = 0.5; households and schools have the lowest importance, *s*^*H*^ = *s*^*S*^ = 0.2. Here, we set low importance for school thanks to the availability of online schooling. We also consider the simplified scenario in which the relative importance of all locations is identical. Therefore, in total, we consider three distinct measures of the COVID-19 epidemic and two assumptions about the relative importance of locations, so in total, we consider six scenarios, as illustrated by Table 4.

**Table 4.** Effective NPIs for each of the six scenarios.

| Scenario | Social function score weights | Status | Results |
| --- | --- | --- | --- |
| TOI | $s^S = s^H = s^W = s^P = 1$ | $I^{as} + I^m + I^s$ | $M = M^H + 0.8M^W + M^S + 0.1M^P$ |
| SOI | $s^S = s^H = s^W = s^P = 1$ | $I^s$ | $M = M^H + 0.8M^W + M^S + 0.1M^P$ |
| DOI | $s^S = s^H = s^W = s^P = 1$ | $D$ | $M = M^H + 0.7M^W + M^S + 0.1M^P$ |
| TOA | $s^S = s^H = 0.2, s^P = 0.5, s^W = 1$ | $I^{as} + I^m + I^s$ | $M = M^H + 0.8M^W + M^S + 0.1M^P$ |
| SOA | $s^S = s^H = 0.2, s^P = 0.5, s^W = 1$ | $I^s$ | $M = M^H + 0.8M^W + 0.9M^S + 0.1M^P$ |
| DOA | $s^S = s^H = 0.2, s^P = 0.5, s^W = 1$ | $D$ | $M = M^H + 0.8M^W + M^S + 0.1M^P$ |

TOI, SOI, DOI stands for identity relative importance of the activities at different locations under three measures of the COVID-19 epidemic: the number of all infections, severe infections, and deceased cases individually. TOA, SOA, DOA stands for intuitive assumptions of the relative importance of the contacts at different locations under three measures of the COVID-19 epidemic: the number of all infections, severe infections, and deceased cases individually.

For intuitive interpretation and simple implementation, we discretize the continuous value of the decay factors into 11 discrete values with equal intervals, 0, 0.1, 0.2, …, 0.9, 1.0. We set the minimal decay factor of the contacts at the public facilities to 0.1 as it is unrealistic to shut down all public facilities. In addition, we assume that *w*^*H*^ = 1, because there is little we can do for the contacts within households. In this way, the total number of solutions is restricted to 11 × 11 × 10 × 1 = 1210 for each of the six scenarios, and an exhaustive search can be used to identify the best NPIs for a certain scenario.

## Results

### Fit the Model from Data

Figure 3 presents the comparison of the simulation results of the proposed A-SEIRD model and the number of confirmed cases in NYC (27). Note that the simulation of the A-SEIRD model obtains the onset of COVID-19. The actual confirmation of COVID-19 infection usually has a six-days-delay (38). Thus, we incorporated the six-days-delay in presenting the simulation results in Figure 3. In general, the simulation results (blue curve) capture the trend of the confirmed case data (black bars). The simulated curve is higher than the confirmed curve, which is common given the wide existence of undiagnosed cases (39). The implementations of the control measures are represented by the red lines, and the four-phases reopening policies are represented by the brown bars. The effects of control measures became obvious with a lag of several weeks, due to the incubation period of the disease. Additionally, the reopening slowed down the decreasing trend.

**Figure 3:**
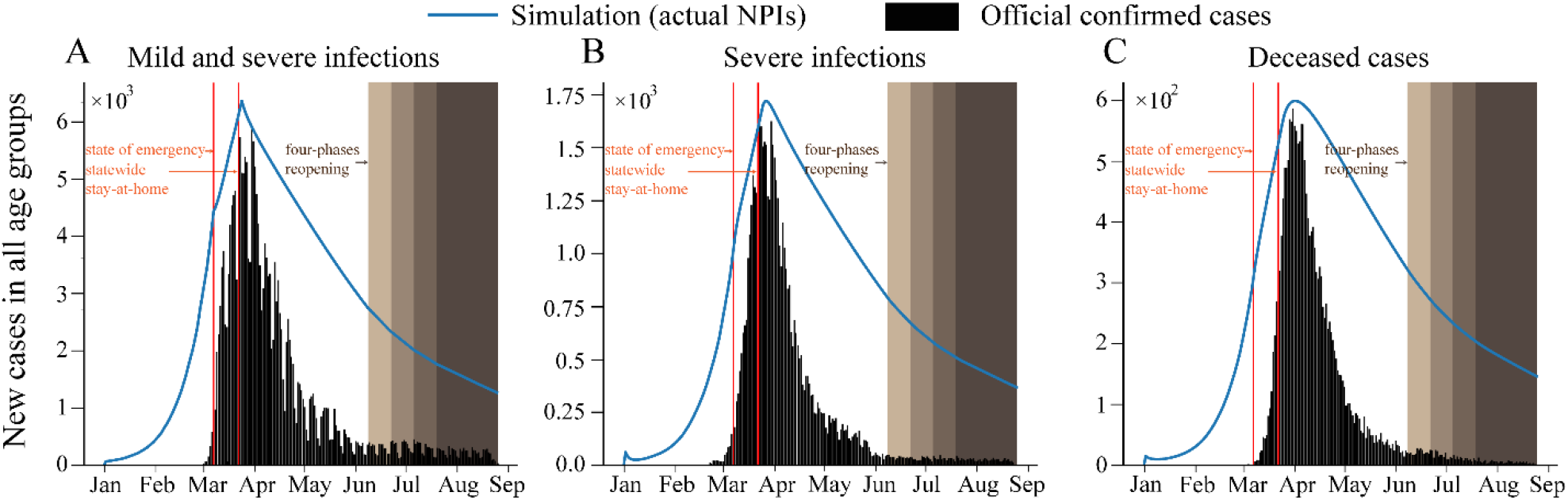
Comparison of our simulation results with the official confirmed case report. (**A**) The number of new mild and severe infections. (**B**) The number of new severe infections. (**C**) The number of new deceased cases. The blue curve and black bars represent the simulated results and the confirmed cases, respectively. The red vertical lines and brown bars indicate the control measures and four reopening phases put in place in NYC.

### Quantify the Effectiveness of NPIs

We present the simulated epidemic sizes with the five common NPIs in Figure 4. Without any control, we would expect a peak of asymptomatic and mild infections in early April. The peak of severe infections occurs two-week later due to disease progression. In such a case, 70% (IQR 60-77) of NYC residents would be infected by the end of 2020. The peak of deceased cases is around early May. Comparing the case with actual NPIs and theoretical no intervention scenario, we observe a 72% (IQR 53-95) decline in the total number of infections and a 76% (IQR 58-96) decline in the number of deceased cases. The effects of each NPIs vary across age categories. More specifically, school closure is effective: the total infections and deceased cases are nearly the same as the results of those without any control (theoretical no intervention). School closure slightly reduces (4% (IQR 3-7)) the number of infections among young people (< 25 years), who are not the most vulnerable. On the other hand, the social distancing for the entire population effectively reduces 47% (IQR 33-84) of infections and 51% (IQR 39-86) deceased cases. In addition, social distancing for the elderly could reduce 47% (IQR 46-56) of total infections and 47% (IQR 46-48) deceased cases in the elderly groups (> 64 years), but not so much for other age groups.

**Figure 4:**
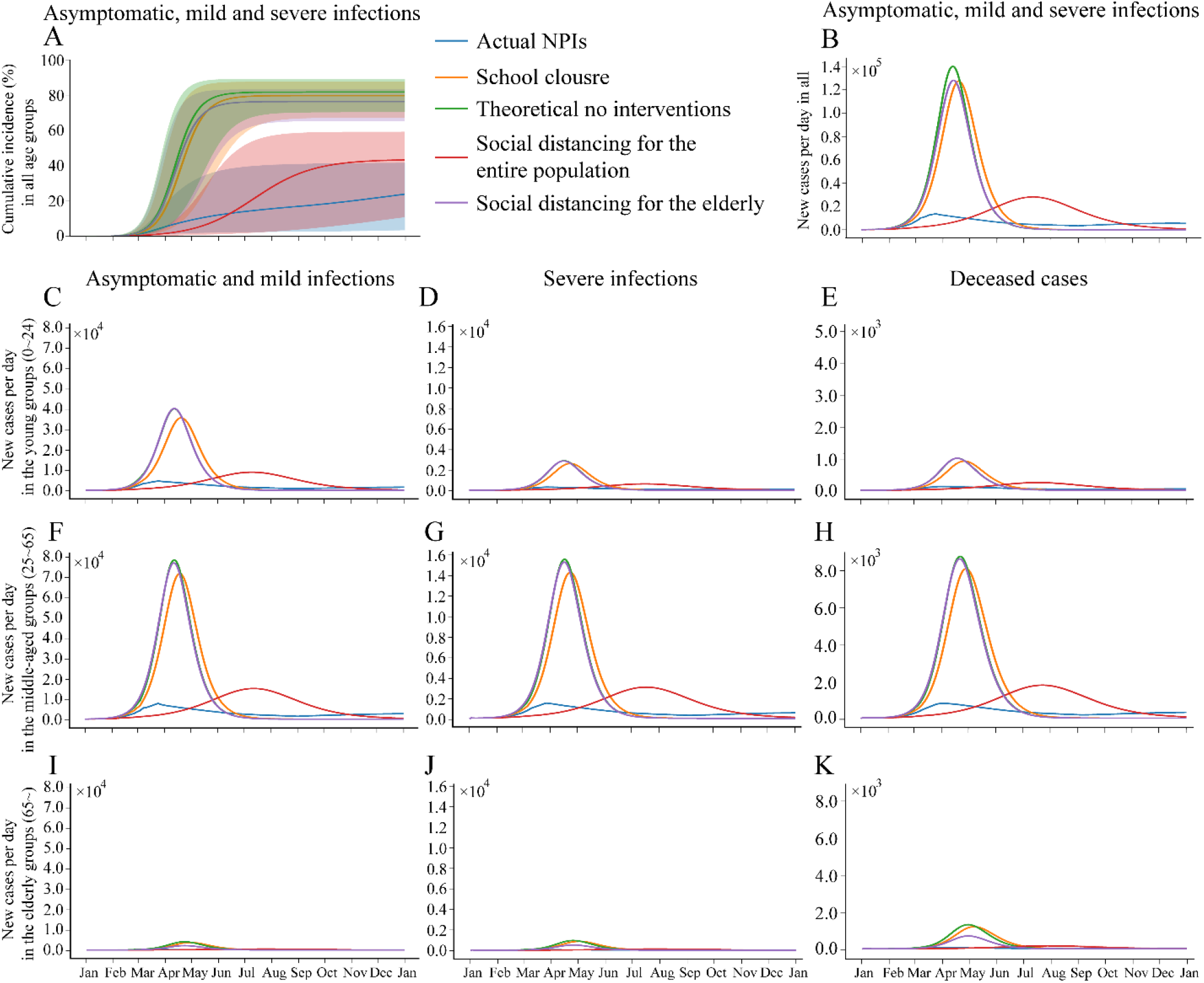
The effects of different NPIs on cumulative cases and new cases per day among age groups. The effects of different NPIs on the cumulative incidence and the number of new cases per day for all are presented in **A** and **B**, respectively. The shaded areas around the coloured lines in panel **A** represent the IQR. The effects of different NPIs on the number of new cases per day for three age groups: the young groups who aged <25 years (**C, D**, and **E**), the middle-aged groups who aged 25 to 64 years (**F, G**, and **H**), the elderly groups who aged > 64 years (**I, J**, and **K**).

We also examined the effectiveness of the adaptive policy, which naturally forms an oscillation curve as shown in Figure 5. The grey blocks represent the periods with stringent strategies (“full lockdown”). We adopt two variants of the adaptive policy from Ferguson et al. (37), “100-severe” (with 100 new severe infections per day as the threshold to trigger the control), and “150-severe” (with 150 as the threshold). Results show that both adaptive policies can reduce the total number of infections and the number of severe infections well. However, the triggered time period of the control is too long, and the interval is only around six days. In practice, such frequent switches between different policies are not realistic.

**Figure 5:**
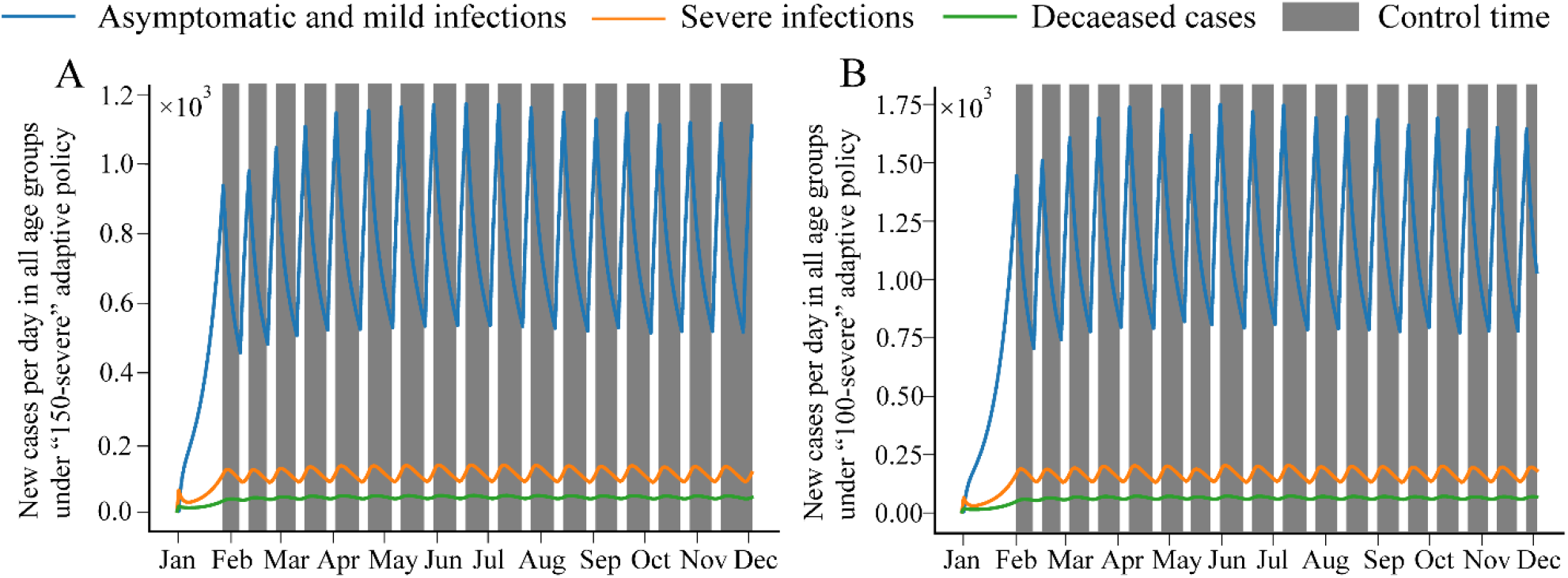
Effects of the two adaptive policies. (**A**) “150-severe” adaptive policy. (**B**) “100-severe” adaptive policy. The blue, orange, and green lines denote the number of new cases per day at all age groups considering the number of asymptomatic, mild, severe infections and deceased cases, respectively. The grey blocks represent the periods with suppression strategies (“full lockdown”).

### Identify the Best NPIs

We derive the best NPIs by exhaustively examining all the possible 1210 solutions for each of the six scenarios introduced in Methods. All scenarios lead to similar NPIs: to reduce the contact intensity at public facilities (social distancing for the entire population) while largely maintaining the function of schools and workplaces. This indicates that, although school closure is associated with lower social disruption, the model still prefers to sacrifice the contact at public facilities because it is associated with higher risk by mixing various age groups in the city.

## Discussion

Since most COVID-19 infections appear to have mild or moderate symptoms that can heal themselves and there are many asymptomatic cases (15), it is critical to reduce the number of severely infected and deceased cases rather than only flattening the total epidemic curve. In this study, we expanded the age-specific SEIR model to develop a new A-SEIRD model that considers the asymptomatic infections of COVID-19 and evaluated the effectiveness of various NPIs in NYC. We found that not all the NPIs were effective, and the social distancing at the public facilities are essential for reducing the number of severe infections and deaths.

Counterintuitively, we find that nearly all the countries decided to close the schools, while our model shows that it is not that effective in controlling the epidemic. As shown in Figure 4, school closure would only slightly reduce the number of total infections compared to the extreme theoretical no intervention policy. As shown in Figure 1. It is obvious that the contacts in schools concentrated on the young population (age <25 years) who make up only a small fraction of the vulnerable population. Meanwhile, the contacts among the young population are also very active in households and public facilities. Thus, school closure alone may not work as effectively as one would have expected. A recent systematic review (40) also concluded that there are no data on the relative contribution of school closures to transmission control and some modelling studies proved that school closure would prevent only 2-4% deceased cases (37). At the same time, school closure would bring more pressure on the medical system since many healthcare workers would be distracted to take care of their children. Jude et al.’s modelling analysis found that 28&8% (95% CI 28&5–29&1) of the health-care workforce need to provide care for children aged 3–12 years (14). A trade-off needs to be taken into consideration while making the school closure decision.

Compared with the social distancing only for the elderly, social distancing for the entire population comes with greater social disruption. Thunström et al. (41) found that the cost of social distancing measures is $7.21 trillion in the USA utilized epidemiological and economic forecasting. Although the social distancing for the elderly would not significantly reduce the total number of infections, it can effectively reduce the number of deceased cases. These results reflect the clinical characteristics of COVID-19: high transmissibility but a high mortality rate only among the elderly (42, 43). That is why the social distancing for the elderly policy has already been adopted in many countries (44).

NPIs lead to social disruptions. We used the proposed A-SEIRD model to identify the NPIs that flatten the curve with minimal social disruptions. We found that the social distancing at public facilities could effectively reduce the number of infections and the number of deaths. As shown in Figure 1, the contacts of the elderly are concentrated in public facilities and households. Although it is difficult to shut down all the public facilities, we suggest that the social distancing for the elderly only at the public facilities would be an effective and practical way to reduce the number of deceased cases without large-scale social disruption. In practice, the protection of the elderly in public areas has already been adopted, such as cresting flow to make room for the elderly in the supermarket.

Apparently, it is not practical to implement the NPIs for a long period, because the social disruption could lead to enormous economic loss, which may, in turn, cause other public health problems. Here, we simulated the ongoing process when relaxing at the beginning of July, August, September, and October as shown in Figure 6. Our model suggests that relaxation after the peak of the outbreak would cause an instant rebound. At the same time, it can be observed that the later relaxing implemented, the smaller the rebound size is. Thus, the resumption of work and production shall be carefully taken place. The adaptive policy, which seems to work in controlling the epidemic size, but leads to unrealistically frequent switches between lockdown and relaxation, making it difficult to be realized (Figure 5).

**Fig 6:**
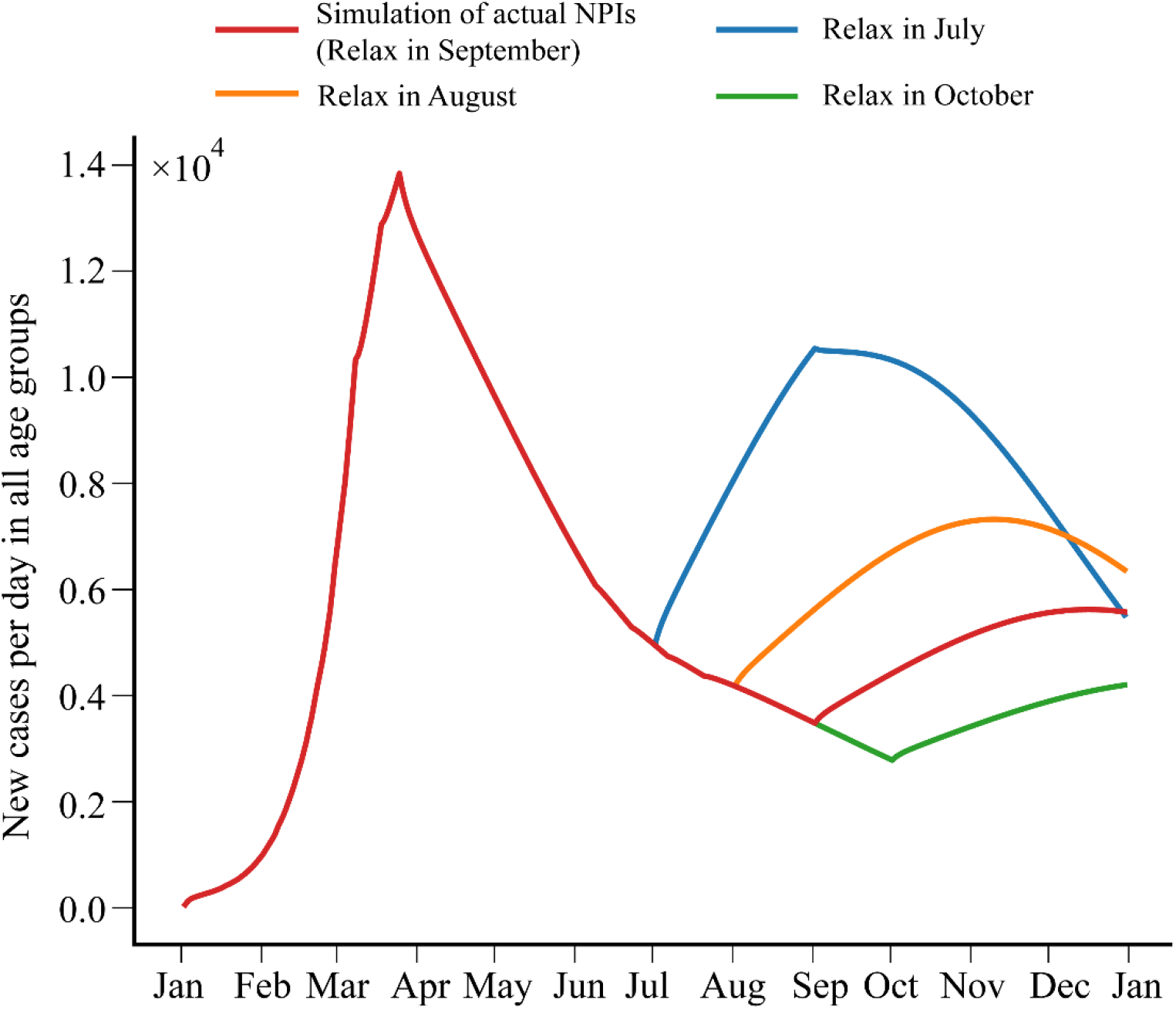
The impacts of policy relaxations in July, August, September, and October. The lines denote the number of new cases. The colours of the lines represent the results of different policies: the simulation of actual NPIs (relax in September) (red), relax in July (blue), August (orange), and October (green).

In conclusion, our study proposed a comprehensive mathematical model that takes into account both the age-specific contacts between people, the latest epidemiological parameters of the COVID-19, and the social disruption. The simulation results provide novel insights into the implementation of NPIs with minimal social disruption. We call for more research on the trade-off between epidemic control and economic loss.

## Code Availability

The source codes have been made available on Github: https://github.com/JasonJYang/Age-specific-SEIRD.

